# Accelerating Exploratory Clinical Research: An LLM-Powered Framework for Cross-Study Data Harmonization and Natural Language Querying

**DOI:** 10.64898/2026.03.04.26345603

**Authors:** Aditya Garg, Arindam Sett, Bethany Baumann, Todd Fry, Sunil Hegde, Bhumika Kapadia, Yogesh Pandit

## Abstract

Clinical research depends on high quality data that is standardized, accessible and interoperable. Yet evolving data standards over time and variations in their implementation hinder the secondary use of clinical trial datasets. Although individual studies adhere to clinical data standards set forth by CDISC (Clinical Data Interchange Standards Consortium), differences in study design, interpretation of complex models, controlled terminologies, and historical conventions create inconsistencies that limit interoperability and complicate cross-study analysis. As a result, harmonizing SDTM datasets across studies is essential to enable efficient secondary use and accelerate evidence generation. To address these challenges, we introduce a framework that leverages Large Language Models (LLMs) to automate the harmonization of study-specific clinical trial data, available in CDISC Study Data Tabulation Model (SDTM) format, into data that is harmonized across trials at scale and also enables natural language querying via a text-to-SQL agent. This system transforms siloed study clinical datasets into interoperable, analysis-ready formats while empowering users to retrieve insights across trials without needing SQL or domain-specific schema knowledge. By constructing a semantic layer and applying retrieval-augmented prompting to models like GPT-4o, our approach improves data access, query accuracy, and scalability across use cases. This work demonstrates the potential of LLMs to transform clinical data workflows, cutting manual effort, substantially reducing manual harmonization effort and query latency in secondary analysis workflows, and enabling faster exploratory analysis and hypothesis generation in clinical research.

## Introduction

Clinical trial data is rich yet fragmented, characterized by heterogeneity across studies, evolving collection instruments, and inconsistent use of controlled terminology. These complexities slow secondary use of data and across study analysis and hinder rapid translation of findings into decisions. To address these challenges, our work pursues two complementary aims: first, to harmonize siloed trial data into interoperable datasets that preserve scientific context across studies; and second, to democratize access by enabling researchers to pose natural language questions that compile into valid SQL against a governed warehouse.

Individual studies typically adhere to the Study Data Tabulation Model (SDTM), and in some cases the Analysis Data Model (ADaM), particularly for regulatory submissions, as required by CDISC and agencies such as the U.S. Food and Drug Administration (FDA). However, novel study design, evolving standards, interpretation of complex models, and historical conventions create inconsistencies that limit interoperability and complicate cross-study analysis. As a result, harmonization within and across trials is essential. By standardizing data for consistency and compatibility, harmonization simplifies interpretation and accelerates analysis. Without it, data remain non-interoperable, prolonging cycle times and limiting the ability to generate insights that drive patient care.

Our approach emphasizes practicality. The system integrates with existing archives and data anonymization services. For data harmonization, it uses CDISC standards, harmonizing values to standard terminologies and moving data based on current SDTM models. It employs a rules-based engine, which is augmented with LLM-based mappings for cases where rules alone are unscalable. Data is then surfaced through a semantic-layer-aware agent that generates SQL with built-in guardrails, returning answers with traceable lineage to the underlying tables and columns. This architecture provides an auditable and iterative framework suited for secondary use of anonymized data. This framework is designed to support exploratory and secondary use of anonymized clinical trial data rather than regulatory submission or decision-critical workflows.

## Objectives

1. Harmonize clinical trial data across trials, enhancing both consistency and analytical insight.
2. Democratize data usage through text-to-SQL conversion, allowing researchers to query databases in natural language without needing knowledge of the underlying schema or SQL syntax.

To achieve these objectives, we begin by examining the nature of clinical data, its formal definition by CDISC, and the reasons it remains inconsistent and in need of harmonization. We then describe the harmonization process itself and how the resulting datasets support text-to-SQL conversion.

Text-to-SQL refers to converting natural language queries into structured SQL statements that can be executed directly on a database. This process democratizes access by eliminating the barrier of technical query languages. Within our framework, the task is divided into four stages:

1. *Semantic Layer Preparation:* constructing a semantic layer that includes table and column definitions, sampled unique values, data types, and joinable columns.
2. *Query Preprocessing:* normalizing natural language queries to ensure they are ready for SQL generation.
3. *SQL Generation:* leveraging LLMs, such as `GPT-4o`, to generate SQL statements from the preprocessed queries.
4. *Query Execution:* executing the generated SQL with tool-assisted validation and self-checking for errors.

While commercial LLMs like GPT-4o possess general biomedical and tabular reasoning capabilities; CDISC-specific semantics are introduced through retrieval-augmented prompting rather than model pretraining, to provide relevant knowledge, preprocessing into a semantic layer remains critical to ensure accuracy and efficiency. The paper therefore provides a detailed examination of harmonization and each stage of the text-to-SQL process, and discusses the potential of LLMs to transform clinical research by making data more accessible, interoperable, and actionable.

## Related Work

The effective analysis of clinical trial data is a critical component of medical advancement. However, this process is frequently hindered by the complexity and inconsistent structure of the data collected. To address this, standardization frameworks from the Clinical Data Interchange Standards Consortium (CDISC), such as the Study Data Tabulation Model (SDTM [5]) and Analysis Data Model (ADAM [4]), have been established to ensure data consistency and facilitate regulatory submissions. Despite the use of these standards within studies, when data is stacked for cross- study analysis there are inherent inconsistencies across studies. This highlights a critical need for more efficient, scalable and automated harmonization methods that allow for reuse of clinical trials data across studies.

Recent advancements in Large Language Models (LLMs) present a transformative opportunity to address these long-standing challenges. The potential for LLMs in the clinical trial space is broad, with researchers exploring their use for a range of tasks from data analysis to final reporting [11]. A key area of investigation is the ability of LLMs to create structured information from unstructured or varied data formats. For instance, studies have demonstrated the successful use of LLMs for clinical named entity recognition, extracting specific entities like medical problems and treatments from unstructured text through careful prompt engineering [9]. This success in data structuring provides a strong precedent for using LLMs to automate the more complex task of harmonizing entire datasets to standards like SDTM. Practical implementations of SDTM harmonization extend beyond theoretical modeling. Patel (2025) outlines a strategic, three-phase framework to standardize CRFs and automate SDTM mapping using tools like R and Python [15]. Guo (2013) introduces a modular SAS-based approach to program SDTM domains in the absence of CDASH metadata [7]. Earlier efforts like Scocca’s SDTM Programming Toolkit laid the groundwork for reusable, metadata-driven approaches to SDTM generation [17]. Beyond harmonization, a second critical challenge is democratizing access to the resulting structured data. Making complex databases available to researchers who may not have specialized knowledge of SQL is crucial for accelerating insight generation. The development of text-to-SQL agents, which convert natural language questions into executable database queries, is an active area of research. Earlier approaches, such as Seq2SQL introduced supervised learning with reinforcement optimization to align query generation with execution [19]. More recent advances have incorporated grammar-aware decoding [12], execution-guided generation [1], and retrieval-augmented transformer-based models tailored to biomedical databases [6, 8]. Comprehensive surveys by Qin et al. and Liu et al. further outline the rapid evolution of NL2SQL methods in the era of large language models[16, 13]. Work in this domain has led to the development of specialized models like MedT5SQL, a transformer-based LLM designed specifically for text-to-SQL conversion in the healthcare context [14].

While this paper provides a solid foundation in LLM-driven text-to-SQL generation, a comparative perspective on existing NL2SQL models can further contextualize the state of the field.

Direct performance comparison between fine-tuned NL2SQL models (e.g., MedT5SQL) and agentic LLM systems is inherently limited, as these approaches differ in training assumptions, deployment flexibility, and schema generalization requirements. Table 1 is intended to contextualize methodological differences rather than provide benchmark equivalence.

**Table 1.** Comparison of fine-tuned text-to-SQL models.

| Model | Architecture Type | Target Domain | Training Dataset(s) | Notable Features |
| --- | --- | --- | --- | --- |
| Seq2SQL[19] | Pointer-based w/ RL | Generic tabular | WikiSQL | Execution-guided training |
| MedT5SQL[14] | Transformer (T5) | Healthcare-specific | Custom clinical datasets | Biomedical schema alignment |
| STaR-SQL[8] | Chain-of-thought + T5 | Generic | Spider, Spider-DK | Self-taught reasoning, CoT prompting |

While previous work has addressed either data structuring or data querying in isolation, this paper introduces an integrated, end-to-end framework. We leverage the power of LLMs for both the harmonization of complex clinical data at scale and the subsequent querying of that data through a natural language text-to-SQL agent. By addressing the entire workflow, from raw data to actionable insights, our work aims to provide a more comprehensive solution that accelerates clinical research. Standardization via CDISC has driven adoption of SDTM and ADaM, yet mapping at scale remains costly and error-prone. Prior efforts often optimize one side of the pipeline. Some focus on metadata-driven SDTM programming and reusable templates, while others explore text to SQL over normalized schemas. Our contribution is to combine these threads with a semantic layer that brings clinical meaning into the prompt context, producing executable SQL that aligns with CDISC semantics rather than raw schema labels. Table 1 positions our approach relative to representative NL2SQL systems and highlights why a retrieval-grounded, schema-aware agent can be more adaptable than a single fine-tuned model in heterogeneous clinical data landscapes.

## Methods

All evaluations were conducted retrospectively on anonymized internal datasets for research purposes.

### Overall System Architecture

Figure 2 presents the architecture: ingestion from archives and pipelines, harmonization via two complementary paths, consolidation in a central Snowflake warehouse, and a semantic layer service that exposes curated metadata to the query agent. The agent uses retrieval to inject this metadata into prompts, then generates and executes SQL with a self-checking loop. Figure 4 details the agent stages: query preprocessing, context construction from the semantic layer, SQL generation with constrained decoding, execution, validation, and response formatting with citations back to tables and columns.

**Figure 1.**
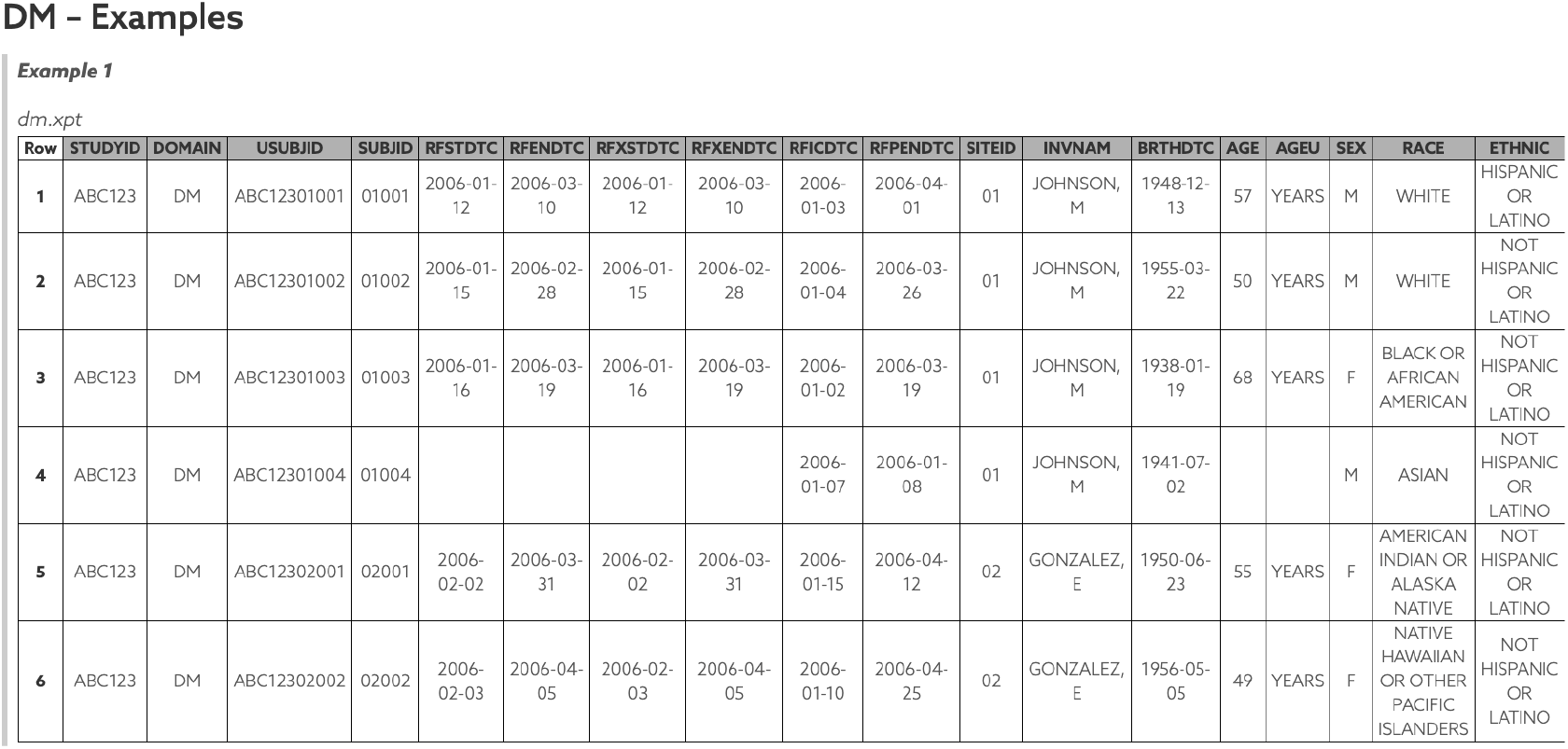
CDISC Demographics (DM) domain synthetic data example [18]

**Figure 2.**
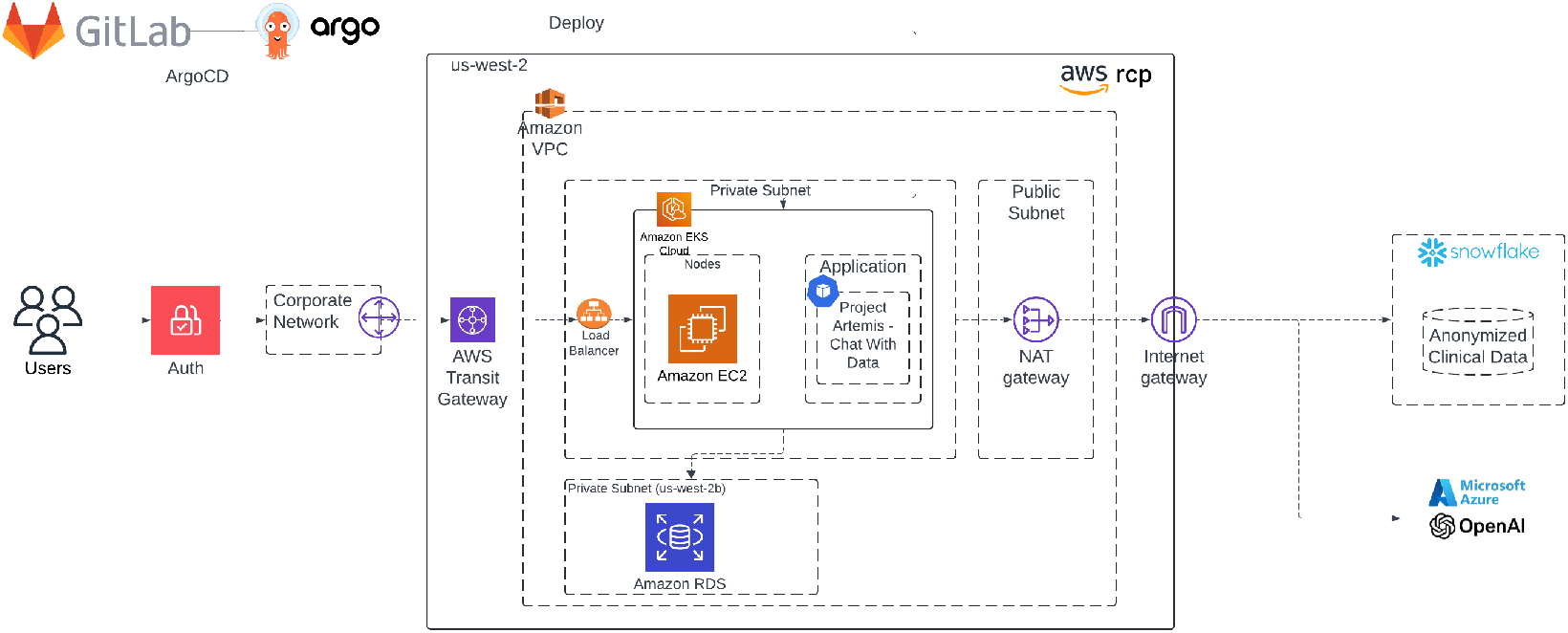
Overall System Architecture

**Figure 3.**
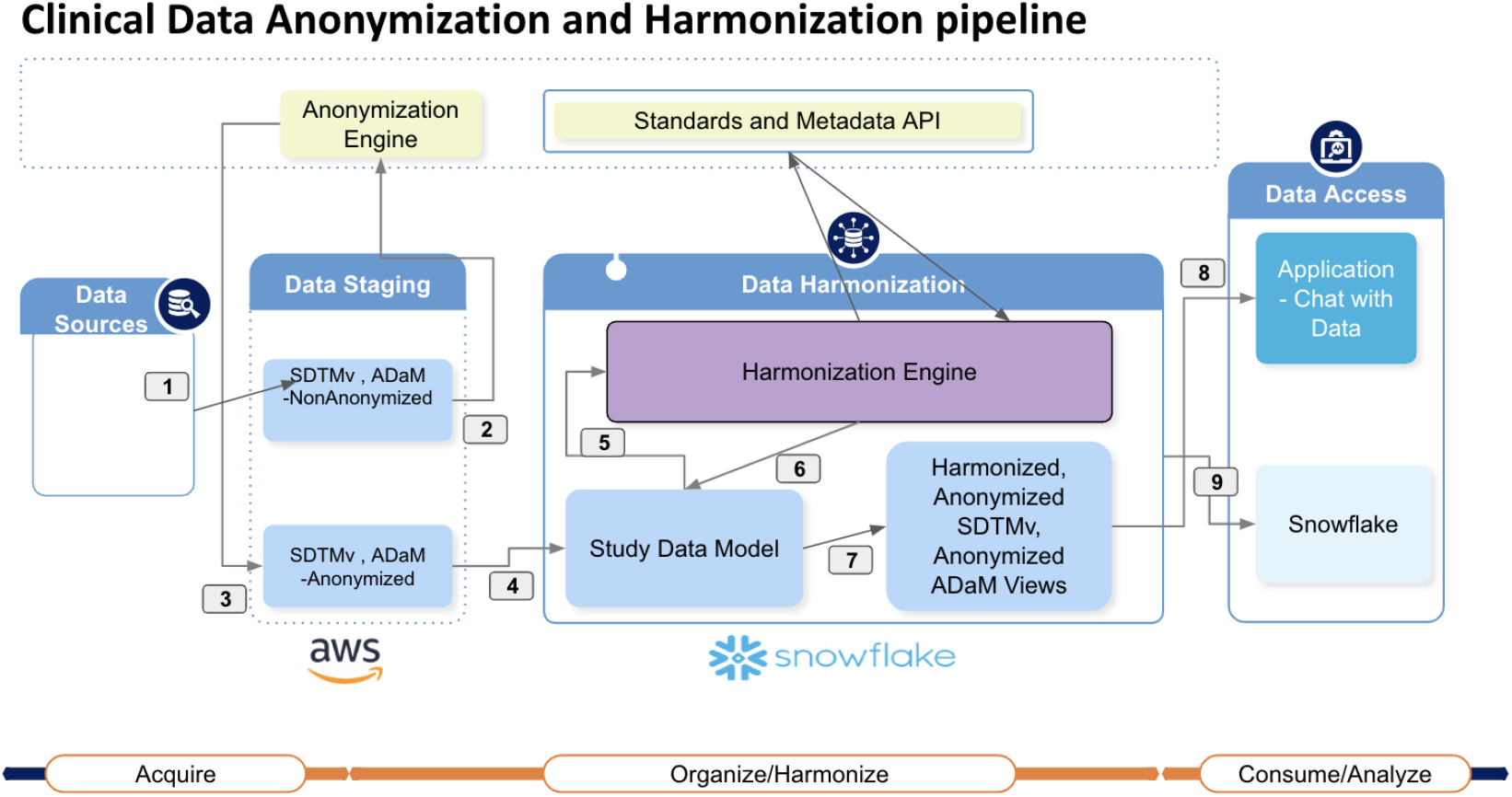
Data Harmonization Workflow

**Figure 4.**
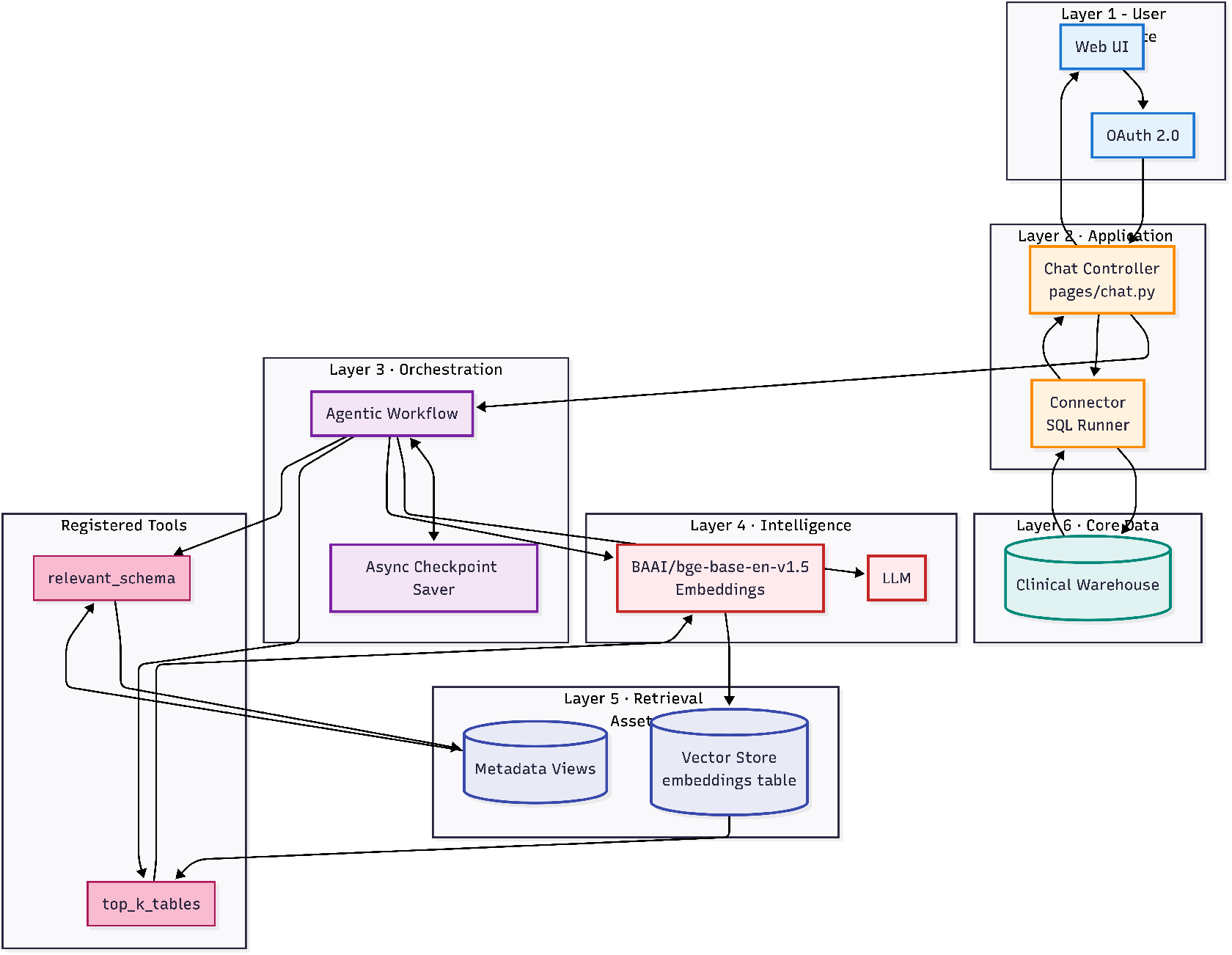
Text-to-SQL Agent Architecture

**Figure 5.**
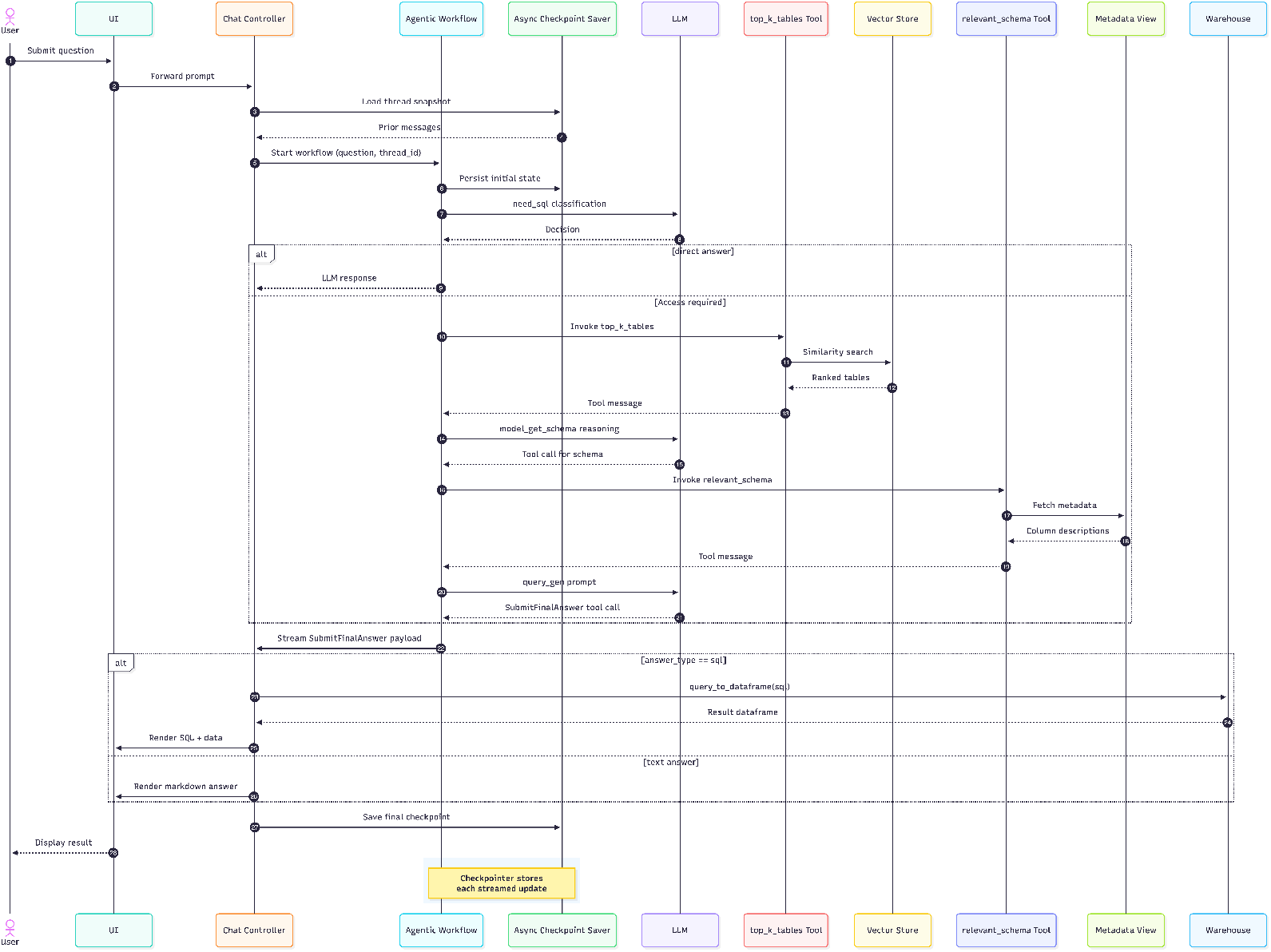
Sequence Diagram for Query Generation and Processing

### Data Harmonization

We automate SDTM mapping via structured metadata and code templates, drawing on methodologies such as Guo’s modular domain programming[7] and Scocca’s toolkit for standardizing domain workflows[17].

### Clinical Data Sources and Preprocessing

Roche/Genentech sponsored clinical trials were the primary source of data. Table 2 provides an overview of the dataset

**Table 2.** Overview of the dataset.

|  |  |
| --- | --- |
| No. of Studies | 511 |
| No. of Domains | 50 |
| No. of Therapeutic Areas | 21 |
| No. of Indications | 113 |

Final Study Data Tabulation Model (SDTM) datasets are ingested from various archives and processing pipelines into secure cloud storage, where a centralized anonymization service applies predefined rulesets to ensure compliance with privacy requirements.

### Cross-Study Harmonization of SDTM-Formatted Data

In this work, harmonization refers not to initial SDTM compliance mapping, but to normalization of SDTM-formatted datasets across studies to address value-level, unit-level, and sponsor-specific inconsistencies. This harmonization process is essential for addressing the inherent complexities of clinical data, which often involves diverse sources, varying formats, and inconsistencies. By standardizing the data, we ensure consistency and compatibility, critical for simplifying analysis and interpretation. This automated approach is designed to overcome the significant bottlenecks associated with manual mapping, which is typically labor-intensive and error-prone.

Harmonization addresses cross-study variation that persists even after SDTM conversions, including unit normalization, controlled terminology alignment, and sponsor-specific conventions. Our rule-based Harmonization Engine uses company standards and a terminology service to standardize anonymized SDTM datasets. It resolves value-level inconsistencies and aligns attributes that drive downstream joins and analyses. When rule coverage is incomplete or ambiguous, we trigger the LLM mapping path that proposes domain and value mappings. Candidate mappings are validated through schema constraints and spot checks against a labeled subset. We record provenance for each transformation and keep both original and harmonized values to support audits. Table 2 summarizes our corpus across 511 studies, 50 domains, and 21 therapeutic areas, which underscores the need for systematic harmonization at scale.

### Text-to-SQL Agent

In our framework, `GPT-4o`serves as a powerful general-purpose reasoning engine, enhanced through retrieval-augmented generation (RAG) and domain-aware few-shot prompting. While `GPT-4o`is not specifically fine-tuned for NL2SQL, we overcome this limitation by injecting structured metadata and curated examples from clinical domains into each prompt. This approach effectively bridges the gap between general LLM capabilities and the structured demands of CDISC-based querying. Compared to domain-specific models, `GPT-4o`offers greater adaptability across varied schemas, while our RAG layer ensures it remains grounded in relevant table definitions, join relationships, and clinical terminology.

### Semantic Layer Preparation

We standardize on text-to-SQL and describe the agent as a composition of retrieval, prompting, constrained generation, and self-check. The semantic layer stores table and column definitions, data types, sampled values, and joinable columns. Retrieval constructs a compact, relevant context per query. SQL generation uses structured prompts with a small set of clinical examples. The self-check step verifies that the query compiles, respects schema and join keys, and returns results within expected cardinalities. If a check fails, the agent revises the query using error-aware hints. This approach reduces hallucination risk and improves latency by avoiding repeated exploratory calls. Figure 4 illustrates this loop.

### Query Generation and Processing

The process of converting a natural language question into an executable SQL query is handled by a multi-step workflow within our agent. First, the user’s natural language query undergoes a preprocessing step that performs lightweight text normalization (e.g., removal of extraneous phrasing and standardization of common expressions) without altering the underlying intent, preparing it for the LLM. Next, the preprocessed query, along with the semantic layer metadata, is passed to a LLM, such as GPT-4o, which is tasked with generating the corresponding SQL statement. This approach leverages the model’s advanced reasoning capabilities to interpret the user’s intent and translate it into formal SQL syntax. Finally, the generated query is executed against the database. A self-checking mechanism is employed to validate the query’s execution and ensure the accuracy of the results, a process that reflects the evolving capabilities of specialized models like MedT5SQL[14] in the healthcare domain. Please refer to the below sequence diagram in 5 for an end-to-end flow.

### Evaluation Metrics

To rigorously evaluate our framework, we assessed the performance of its two core components: data harmonization and the text-to-SQL agent. The effectiveness of the data harmonization module will be measured by its accuracy. For the text-to-SQL agent, performance will be evaluated across three key dimensions: accuracy, consistency, and performance.

### Harmonization Performance

- Coverage Rate
  ‐ The proportion of non-codelist raw data values that were successfully mapped to any entry in the codelist
  (Number of mapped values /Total number of values requiring mapping) × 100

- Speed
  ‐ The time needed to complete harmonization

- Accuracy
  ‐ The proportion of value mappings that are considered correct according to manual review
  ‐ (Number of value mappings considered correct /Total number of mapped values) x 100

Inter-reviewer agreement was not assessed and represents a limitation of the current evaluation. Unmapped values include both true system failures and values that were irrecoverable due to upstream data quality limitations.

### Text-to-SQL Performance

- Accuracy
  ‐ *Query Interpretation Accuracy:* The percentage of natural language queries correctly interpreted and converted into accurate SQL statements.
  *Execution Accuracy (EX):* The percentage of SQL statements that return the correct results when executed against the database [2].

- Consistency
  ‐ *Query Interpretation Consistency:* The percentage of times the agent generates the same SQL query for a given natural language query across multiple attempts.
  ‐ *SQL Execution Consistency (EC):* This metric focuses on the reliability of the output for questions that have at least one correct execution

- Performance
  ‐ *Response Time:* The average time taken by the agent to respond to a query and generate the corresponding SQL statement
  ‐ *Valid Efficiency Score (VES):* A combined evaluation of accuracy and SQL efficiency [2]

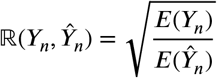

This measures how efficient the generated query is compared to the reference.

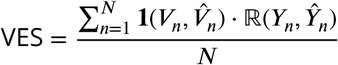

This averages these efficiency ratios only for valid (correct) queries, giving a single score that reflects both accuracy and runtime efficiency.

where:

*N* is Total number of evaluated queries or test cases

*Y*_*n*_ is Ground-truth SQL statement (or reference query) for the n-th test case

*Ŷ* _*n*_ is Model-generated SQL statement for the n-th test case.

*E*(*Y*_*n*_) is Execution cost or runtime efficiency metric of the reference query *Y*_*n*_.

*E*(*Ŷ _n_*) is Execution cost or runtime efficiency metric of the generated query *Ŷ* _*n*_.

ℝ(*Y*_*n*_, *Ŷ* _*n*_) is Relative efficiency ratio between the reference and generated queries; values closer to 1 indicate similar efficiency.

*V*_*n*_ is Expected (ground-truth) output of the reference query.

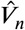 is Output produced by the generated query.

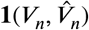 is Indicator function: equals 1 if the generated query output matches the expected output (i.e., correct result), otherwise 0.

‐ *Scalability:* The agent’s ability to handle an increasing number of concurrent queries without significant degradation in performance.

## Results

We evaluated performance across two core components of the framework: data harmonization and the text-to-SQL agent.

### Performance of Data Harmonization

Evidence indicates that automated harmonization is associated with substantial reductions in manual mapping effort and accelerates processing, especially in domains with heterogeneous source data [10].

The performance of the harmonization engine was measured first using coverage rate: the proportion of raw data values that were mapped to any entry in the code list. Here, we present the values for two domains, Demographics (DM) and Laboratory Test Results (LB). We had a 100% coverage rate for the DM domain, except values for RACE. In the case of RACE, many values didn’t match the codelist and were not possible to decipher, as is inherent in the case of mapping clinical data retrospectively. Within the LB domain, the coverage rate was 67.6% across all variables. This is expected as some variables have data quality issues that cannot be corrected and result in data that is incomplete. Coverage rates for variables that span across multiple domains and were harmonized using LLM were also measured. As an example, xxLOC is a variable that describes the anatomical location of a subject and spans across 17 domains. The coverage rate for xxLOC was 95.54%.

The speed of harmonization was measured and as expected was an improvement over manual effort. LLM harmonization for the xxLOC variable could be completed in 100 minutes. This is for a total of 12,502 unique values that required harmonization. The LLM harmonization results were sorted and reviewed manually for corrections in a few hours. Manual harmonization of this variable would not be possible, and if attempted, would take months. Applying pre-programmed and LLM-generated harmonizations was performed through the harmonization engine. Building the engine and harmonizing the raw data with it required 7.5 minutes for DM (122,770 rows of data) and 24 minutes for LB (61,892,517 rows of data).

The accuracy of harmonization was measured by the proportion of value mappings that were considered correct according to a reviewer. For non-LLM harmonized variables the harmonization used pre-programmed logic. A review showed the expected, specified harmonizations were applied, so these mappings executed as specified by predefined logic and were not observed to contain errors during review. For LLM-harmonizations, a reviewer checked the LLM output for low quality harmonizations, including those that were not from the codelist, incorrect mappings, or had no value assigned. For the xxLOC variable, 2% of the LLM output harmonization values were corrected by the reviewer. This gives an accuracy by our metric of 98%. We additionally retain the original values alongside the LLM harmonized values for the end user, so that they can transparently interpret the accuracy of the LLM harmonized values.

### Performance of Text-to-SQL Agent

The following evaluation represents a pilot-scale assessment intended to validate feasibility rather than establish benchmark-level performance. Evaluation was conducted on a corpus of 22 natural language questions authored by domain experts, each paired with a reference SQL query. Each natural language question was paired with a single reference SQL query, acknowledging that multiple semantically equivalent formulations may exist. Both a custom semantic-layer-aware agent and a prebuilt LangChain [3] text-to-SQL agent were tested.

Table 3 shows the Execution Accuracy (EX), Valid Efficiency Score (VES), and mean end-to-end agent latency across 3-fold cross-validation. Overall performance metrics reported in Table 3 are computed across all queries, independent of difficulty tier. Accuracy profiles are shown in Figure 6; where execution consistency is computed over questions with at least one correct execution. The prebuilt agent evaluates using only database-resident information, while the custom agent augments prompts with a semantic layer comprising table/column definitions, sampled values, and explicitly defined joinable columns.

**Table 3.** Performance of text-to-SQL agent from top 3 runs.

| Trial | Semantic Layer Aware Agent |  |  | Prebuilt LangChain Agent |  |  |
| --- | --- | --- | --- | --- | --- | --- |
|  | EX (%) | VES (%) | Latency | EX (%) | VES (%) | Latency |
| 1 | 63.63 | 63.61 | 12.324 ± 2.621 | 13.64 | 16.25 | 53.65 ± 25.20 |
| 2 | 72.72 | 79.26 | 13.05 ± 2.44 | 9.09 | 11.15 | 50.58 ± 5.58 |
| 3 | 72.72 | 72.02 | 10.50 ± 1.72 | 13.64 | 15.85 | 63.06 ± 58.73 |
| Across-trial Mean | **69.69** | **71.63** | **11.96** | 12.12 | 14.42 | 55.77 |

**Figure 6.**
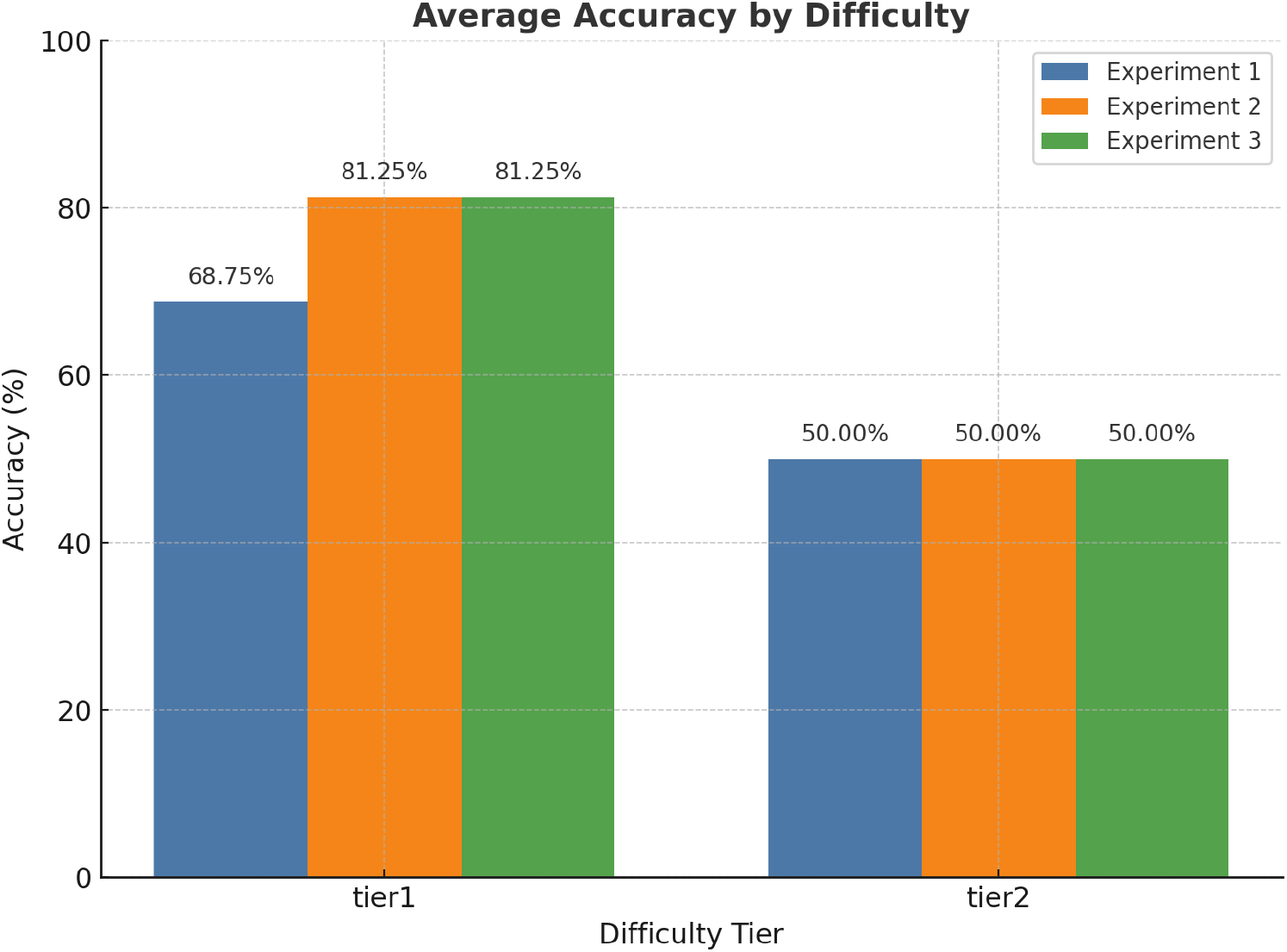
Accuracy Stratified by Query Structural Complexity

Query difficulty tiers were defined a priori based on structural properties of the reference SQL, independent of model performance

- Tier 1 (Single-domain queries)
  ‐ ≤ 1 join
  ‐ Single SDTM domain
  ‐ No nested subqueries

- Tier 2 (Multi-domain queries)
  ‐ ≥ 2 joins
  ‐ Multiple SDTM domains
  ‐ Value-level filters across domains

Across three independent trials, the semantic-layer-aware agent consistently outperformed the baseline. Mean execution accuracy reached nearly 70%, compared to 12% for the schema-only baseline. Similarly, the Valid Efficiency Score (a joint measure of accuracy and query efficiency) averaged 71.6% for the semantic-layer agent versus 14.4% for the baseline. Latency was also markedly improved: 11.9 seconds on average compared to 55.8 seconds. These gains were particularly pronounced in multi-table queries requiring complex joins and value-level filters, where semantic context is critical.

## Discussion

These results demonstrate the importance of embedding structured metadata into LLM prompting. By grounding GPT-4o in curated table and column definitions, join relationships, and sampled values, the semantic layer substantially improves both correctness and stability. The benefits are most evident in queries involving multiple domains, where schema-level information alone is insufficient.

### Potential Applications

#### Some examples of use-cases are

- Analysis of Clinical Safety Data - Access to clinical safety parameters such as adverse events, drug exposure, lab assessments, and biomarker data matched with patient metadata to trans late preclinical findings and develop toxicity models.
- Access to clinical data for enrolled patients to correlate immune responses with safety signals and other treatment-related factors for comprehensive pharmacodynamic (PD) analysis.
- Data Integration for Modeling - Ability to combine clinical data with other data modalities, such as PK/PD, genetics, biomarkers, with additional endpoints for efficient modeling efforts.
- Link clinical data with high-dimensional data for translational biology.
- General queries that span across studies for further analysis and complex exploration.

### Interpretation of Key Findings

The semantic-layer approach yields a substantial improvement within the evaluated exploratory workload, suggesting that the “make data right” (harmonization) and “make data usable” (natural language access) problems can be addressed synergistically. By lowering the technical barrier for querying and accelerating analysis cycle times, this framework has the potential to broaden the use of clinical trial data across research groups, improving decision-making and ultimately supporting faster exploratory analysis and hypothesis generation in clinical research.

### Governance and Compliance

A central requirement in clinical data systems is traceability. Our framework enforces governance through anonymized datasets, role-based schema access, and full logging of prompts, retrieved context, generated SQL, execution results, and user approvals. Updates to harmonization rules and semantic layers undergo review and change control, ensuring efficient AI-powered usage. Monitoring dashboards track execution errors, schema violations, and unusual query patterns to maintain oversight.

### Limitations

Several limitations remain. First, the framework depends on the accuracy and freshness of the semantic layer. Shifts in domain conventions or outdated metadata can reduce effectiveness. Second, ambiguity in natural language queries can still yield plausible but incorrect SQL, particularly when user intent is underspecified. Third, execution accuracy, while important, does not guarantee clinical validity; generated results may still require domain expert review in high-stakes contexts. Evaluation was conducted on a limited number of expert-authored queries and may not reflect broader user behavior or query complexity.

Even though this is a clinical data system, it is not intended for GxP-regulated use (i.e., it is not used to generate, process, or manage data that supports regulated decisions such as clinical trial submission, patient safety determinations, product release, or other compliance-critical outcomes). Because the intended use is non-GxP, formal GxP validation artifacts (e.g., full traceability matrix as part of CSV/CSA, IQ/OQ/PQ packages, etc.) are not required.

### Future Directions

To address these limitations, several extensions are planned

- Scaling the evaluation corpus to include a wider range of question types and domains, increasing robustness.
- Integrating declarative data tests and statistical drift monitoring to detect inconsistencies across studies.
- Establishing active feedback loops where users rate outputs, generating automated updates to semantic layers and harmonization rules.
- Implementing tiered validation, combining automated constraint checks, statistical monitoring, and human review for critical domains.

These enhancements will improve reliability, maintain trust in system outputs, and ensure scalability across diverse clinical research contexts.

## Conclusion

This work presents an integrated framework that couples automated harmonization with semantic-layer-aware natural language querying. By leveraging both rule-based standards and LLM-assisted mapping, we create interoperable datasets that can be explored through text-to-SQL without requiring specialized technical skills.

Early results show substantial improvements in accuracy, latency, and scalability over baseline approaches, supporting the vision of faster, more reliable clinical data analysis. As the system matures, it offers a practical approach for improving accessibility and efficiency of exploratory clinical data analysis, enabling broader access to clinical trial data, and supporting more efficient exploratory clinical data analysis.

## Data Availability

All data produced are available online at ClinicalTrials.gov

